# Stroboscopic Light Stimulation Safety Within and Beyond Laboratory Settings: Observational Evidence and Practical Guidance

**DOI:** 10.1101/2025.11.17.25340398

**Authors:** David J. Schwartzman, Trevor Hewitt, Timo T. Schmidt, Josemir W. Sander, Danny Nacker, James Thurbon, Vince Polito, Xaver Funk, Fiona Macpherson, Tom Galea, Gavin Lawson, Jimi Simpson, Romy Beauté, Anil K. Seth

## Abstract

Stroboscopic light stimulation (SLS) reliably evokes geometric visual hallucinations and, in some contexts, altered-state experiences. Its increasing use in research, public, recreational and exploratory clinical settings creates a need for proportionate safety guidance. The main concern is a visually triggered seizure in a susceptible individual; reported non-epileptic responses include discomfort or distress. We combined a retrospective survey of safety practices and reported events across four laboratories, provider-reported operational data from two commercial SLS platforms, and a focused review of photosensitivity and sensory intolerance. Laboratory contributors reported 20 non-serious events that interrupted or modified participation, with no reported seizures or syncopal episodes. One commercial provider reported nine medically important or otherwise notable events across approximately 3.8 million sessions. The other reported five such events across more than 400,000 sessions. Because commercial adverse-event capture was passive and the laboratory protocols and monitoring procedures were heterogeneous, these figures cannot be used to estimate incidence or establish screening effectiveness. This synthesis informed a provisional Sussex Strobe Safety Screening Questionnaire (4SQ) and a layered, context-specific risk-management approach. The resulting framework distinguishes non-serious tolerability concerns from the principal medical risk of a visually provoked seizure in a susceptible individual, and supports conservative monitored familiarisation exposure, immediate stopping procedures and systematic adverse-event surveillance. The predictive performance of the 4SQ and the effectiveness of these precautions require formal evaluation; absolute SLS risk remains uncertain.

## 1.0 Introduction

Stroboscopic light stimulation (SLS), usually delivered to closed eyes at frequencies between ∼5–50 Hz, evokes stroboscopically induced visual hallucinations (SIVHs) comprising dynamic geometric patterns and colours. Occasionally, more complex visual hallucinations, such as faces, people or scenes, are reported, as are altered-state experiences ^1–3^. Comparable phenomena occur with open-eye stimulation using diffusion goggles ^4^. As SLS use expands beyond specialist research laboratories, clear and proportionate safety frameworks are required.

Contemporary SLS research has addressed multiple, partly overlapping aims, including investigation of the neural correlates of SIVHs using fMRI and EEG, particularly in relation to neural entrainment and thalamocortical dynamics ^1,5–13^. These studies often combine electrophysiological and phenomenological measures to relate neural dynamics to subjective visual experience. SLS has also been used to examine altered states of consciousness (ASCs; ^2,11,14,15^) and to model simple visual hallucinations ^7,16,17^.

Interest has grown in the potential therapeutic applications of SLS, particularly at gamma (∼40 Hz) and alpha (9–11 Hz) frequencies, for conditions such as Alzheimer’s disease and depression ^18–21^. However, the evidence for therapeutic benefit remains preliminary and inconclusive ^18,22^.

Against this backdrop, we synthesise cross-laboratory safety practice, retrospective reported-event data and operational reports from two commercial SLS providers, alongside a focused review of photosensitivity and sensory intolerance. Our aims are to describe current practice, identify recurrent tolerability and safety considerations, and develop a provisional, evidence-informed screening checklist for formal evaluation. The available data characterise reported adverse responses and recurrent safety considerations but cannot establish population incidence or screening effectiveness.

### 1.1 Medical risks and sensitivities associated with SLS

#### Primary risk factor: photosensitive seizure

The main medical concern associated with SLS is a visually provoked seizure in an individual with photosensitivity. Photosensitive epilepsy (PSE) refers to epilepsy in which seizures can be provoked by visual stimuli. A photoparoxysmal response (PPR) is an epileptiform electroencephalographic response to visual stimulation; it indicates photosensitivity but is not itself a clinical seizure or an epilepsy diagnosis ^23,24^. Photosensitivity is more common in younger people ^23^ and in particular epilepsy syndromes ^24^, but can occasionally persist or present later in life ^23,52^. Photophobia instead refers to discomfort or intolerance associated with light and should not be treated as synonymous with PSE ^35^. These distinctions are important because the present synthesis includes medical events and non-serious tolerability outcomes.

Factors mainly relevant to visually provoked seizures or seizure susceptibility include a personal history of epilepsy or seizures, a first-degree family history of photosensitive epilepsy, or younger age. Photosensitive seizures are heritable ^24^, and genetic susceptibility contributes broadly to epilepsy ^25^. Other factors may include sleep deprivation, recent alcohol or recreational drug exposure, acute illness, and some medication or withdrawal contexts. These factors do not contribute equally and should not be combined into a single risk score. PSE is significantly more common in children and adolescents than in adults ^23^, although later-life photosensitivity has also been documented ^52^.

Sleep deprivation can increase seizure likelihood in people with epilepsy ^26,27^. Reduced sleep can increase cortical excitability and epileptiform discharges, particularly in idiopathic generalised epilepsies ^26–29^, while diary-based and longitudinal studies have reported increased seizure risk after acute sleep loss ^30,31^. Medication-related risk is instead drug-, dose- and context-specific and may be concentrated during toxicity, recent dose change, withdrawal, interacting illness or combinations of pro-convulsant factors.

Pregnancy involves physiological, hormonal and psychological changes that may influence seizure control across trimesters ^32^. Altered drug metabolism, disrupted sleep, stress and fatigue may further affect seizure susceptibility during pregnancy. Because direct evidence on SLS during pregnancy is lacking, pregnancy should be treated as a precautionary exclusion in research protocols rather than as an established SLS-specific risk factor.

#### Secondary risk factors: sensory sensitivities and photophobia

Separate from seizure susceptibility, anxiety, migraine or photophobia, sensory sensitivity, and active psychiatric symptoms may affect the tolerability of SLS. Evidence linking these characteristics to serious SLS-related medical events is limited; they are mainly potential markers of discomfort, distress or early withdrawal rather than established seizure predictors.

Below, we summarise these sensitivities along with their prevalence.

##### Anxiety

In the UK, medical consultations for anxiety have recently increased markedly ^33^, with prevalence estimates ranging from 3.8% to 25% of the population ^34^. Anxiety and panic disorders have been associated with reduced tolerance to light ^35^, while flicker can evoke visual discomfort ^36^. These observations support treating anxiety as a potential tolerability factor; they do not establish anxiety as a predictor of serious SLS events.

##### Migraine

Migraine affects approximately 14–15% of people globally and accounts for about 4.9% of years lived with disability ^37^. Photophobia is common; bright or flickering light is frequently reported as a trigger, although apparent triggering may sometimes reflect photophobia during the premonitory phase ^35,38^. High-contrast SLS may therefore be less tolerable for some people with migraine.

##### ASD

NICE estimates that autism affects around 1.1% of UK adults ^39^. Sensory hypersensitivity and heightened physiological responses to sensory stressors have been described in studies of autistic children ^40,41^, although these features are heterogeneous and not universal. In this adult framework, the relevant screening target is self-reported visual or sensory intolerance, not the diagnosis itself.

##### ADHD

In an online survey, photophobia was self-reported by 69% of adults with ADHD or ADHD symptoms and 28% of respondents without ADHD ^42^. ADHD itself is not a contraindication; the relevant consideration is self-reported light discomfort or sensory intolerance.

##### Psychosis

Global estimates place the prevalence of schizophrenia at around 0.3%, although estimates vary by method ^43,44^. Because SLS evokes vivid visual phenomena, active psychotic symptoms may plausibly increase distress or confusion; this is a precautionary inference rather than evidence of an SLS-specific event rate.

### 1.2 Public incidents shaping perceived risk and regulation

Several widely reported incidents have shaped risk perception and regulation of flashing visual stimuli. The 1997 “Pokémon incident” in Japan involved rapidly alternating red-blue flashes at approximately 12 Hz, followed by seizures and other symptoms in many viewers, including some children without a previous epilepsy diagnosis ^45^. Japan subsequently introduced broadcasting restrictions on provocative flashes and saturated-red flicker ^45^. The London 2012 Olympics’ promotional footage was withdrawn following public reports of seizures associated with the animation, although these reports did not medically verify these events ^47^. These broadcast and public-event examples establish the importance of provocative stimulus characteristics, but are not directly comparable with screened, closed-eye laboratory SLS.

### 1.3 Current screening practice

Screening protocols vary widely in context, scope, design, and their evidence base. Laboratory studies typically use bespoke self-report questionnaires to exclude known PSE and other neurological or psychiatric vulnerabilities. In contrast, public installations often rely on simplified health declarations or verbal briefings with varying degrees of formality. Both approaches have limitations, as many focus narrowly on epilepsy, overlooking other clinically relevant sensitivities. Protocols are also context-specific and often insufficiently documented for replication or cross-study comparison.

To address these gaps, we developed the Sussex Strobe Safety Screening Questionnaire (4SQ) as a provisional, evidence-informed, non-diagnostic checklist designed to support structured risk review and proportionate precautions. It integrates neurological, psychiatric, sensory, medical, substance, sleep and medication considerations, but its predictive performance and effect on adverse events will require formal evaluation.

## 2.0 Methods

### Ethics, consent and data governance

This study was a secondary analysis of anonymised, non-identifiable summary information supplied by contributing laboratories and commercial providers. We did not recruit participants for this synthesis and did not receive or access identifiable participant-level data. Under University of Sussex institutional guidance, analysis of these non-identifiable secondary data did not require new research ethics review or a separate participant-consent process. Participants in the contributing academic studies provided informed consent before taking part under the relevant local ethics procedures; depending on the study format, consent was recorded in writing or electronically. Commercial providers supplied aggregate, non-identifiable operational reports rather than participant-level research data. Their users were not recruited as research participants in this synthesis, and the providers did not seek research consent from them for this secondary analysis. Any service-use safety acknowledgements obtained by the providers were not treated as research consent.

### 2.1 Cross-laboratory and commercial data acquisition on SLS safety and practice

On 24 July 2025, a structured Qualtrics portal was distributed to four SLS research groups and two commercial providers with substantial SLS experience; the data were accessed 28 days later, on 21 August 2025. It requested the contributor’s identity and reporting scope; reporting period; number and purpose of studies or sessions; device, eye state, frequency, brightness and duty cycle; participant or user counts; screening and risk-management procedures; and event narratives or supporting documents. A major event was defined as a seizure or another negative outcome with medical implications, and a non-serious event as a negative outcome that obstructed or stopped participation without reported medical implications. Contributors retrospectively determined which events met these definitions. Event descriptions were supplied by the contributing teams and were not independently confirmed against medical records. For online studies, follow-up was limited to information available to the original research teams. Narratives were grouped post hoc into descriptive symptom categories, using the primary stated reason for stopping when several symptoms were reported. Because populations, protocols, exposure units and surveillance procedures differed, we report sources separately and do not treat the synthesis as a meta-analysis.

#### Research laboratories

Four research laboratories contributed data from studies conducted between 2020 and 2025 in the UK, Germany, the Netherlands and Australia. The studies involved mainly healthy participants, alongside adults reporting depressive symptoms. Contributor summaries described one study from Lab A (reported N = 20), twelve from Lab B (N = 461), six from Lab C (N = 480) and five from Lab D (N = 109). These are reported study totals rather than verified unique-person denominators: some study-level sample sizes were unavailable, and participation in more than one study could not be excluded. Where available, laboratories provided summary information on screening procedures and adverse-event logging.

##### SLS characteristics

Across laboratories, SLS was delivered using commercial and custom hardware. Devices included commercial stroboscopes (Lucia N003, Light Attendance GmbH, Innsbruck, Austria; Roxiva RX1, roXiva Ltd); custom-built stroboscopes (Lumenate Growth Ltd, Bristol, UK; University of Sussex); a projector (Epson EB-G7400U); and standard computer monitors at maximum brightness.

#### Commercial SLS providers

Data from two commercial SLS providers were obtained. Provider A operates a mobile application delivering closed-eye SLS through a smartphone flashlight, with frequency, brightness and duty cycle modulated in synchrony with audio. It reported approximately 3.8 million sessions from 2021 to 2025 and approximately one million users. As reported sessions substantially outnumber users, repeated use contributes to the session total. The dataset does not establish whether reported events were followed up independently or whether adverse-event capture was complete.

Provider B manufactures a configurable LED SLS device used in individual and group settings with more than 400,000 sessions over approximately five years. This is a minimum reported operational session count rather than a unique-user denominator: the same user could contribute more than one session, and additional consumer use was not included. Event reporting was passive and the completeness of follow-up is unknown. The two providers are described separately.

## 3.0 Results: laboratory-based studies

### 3.1 Laboratory-based SLS survey

Most studies (n = 21) used fixed-frequency SLS with reported frequencies spanning 1–80 Hz; all used a square-wave SLS signal. Stimulation protocols typically used frequencies within the alpha frequency range (8–12 Hz; n = 9), with one laboratory also testing 15 Hz stimulation. A subset (n = 3; total n = 95) used dynamic frequency shifts between 3 and 15 Hz.

Flash duration and inter-flash interval (duty cycle) varied, with most protocols adopting a 50% duty cycle (equal flash and inter-flash interval), with values ranging from 25% to 75%.

Brightness levels varied by device: mean luminous flux for stroboscope-based setups was ∼5,373 lumens (SD = 326); the projector operated at 447.2 cd/m²; monitor setups typically delivered ∼300 cd/m². As luminous flux (lumens) and luminance (cd/m²) quantify different properties, these values are not directly comparable and are reported as provided by each laboratory.

Session duration ranged from brief exposures (8 s) to extended stimulation (≤ 25 min), with some protocols including ramp-up periods and rest intervals.

### 3.2 Participants

Across the laboratory datasets, most studies recruited young adults, predominantly from university or student populations. The two depression studies at Laboratory B recruited a broader adult population reporting depressive symptoms. The commercial providers reported sessions or users rather than research samples and did not provide equivalent demographic profiles. Participants were not counted more than once within any individual study, but some individuals may have participated in more than one contributing study; cross-study deduplication was not available.

### 3.3 SLS Safety Screening Across Laboratories

Screening and risk-management procedures varied across groups, reflecting differences in study populations, aims, and institutional requirements (Table 1). We collated safety protocols from the four laboratories (Labs A-D) and identified a shared minimum screening set, alongside areas of divergence.

**Table 1.** Comparison of screening criteria for SLS across four laboratories (Labs A-D) and two commercial providers (Provider A = app-based; Provider B = device-based). Each row lists a screening criterion, with columns indicating whether it was explicitly included in the available protocol. Symbols: ’@’ criterion included; ’—’ not included; ’@’ qualified or partial inclusion. PSE = photosensitive epilepsy; ’≤3y’ = within the last 3 years; FHx = family history; u/wk = units per week; psych. = psychiatric.

| Exclusion criterion | Lab A | Lab B | Lab C | Lab D | Commercial Provider A | Commercial Provider B |
| --- | --- | --- | --- | --- | --- | --- |
| History of epilepsy or PSE | ✓ | ✓ | ✓ | ✓ | ✓ | ✓ |
| Family history of PSE | ✓ | ✓ | — | — | ✓ | ✓ |
| Photophobia | ✓ | ✓ | ✓ | ✓ | ✓ | — |
| Migraine | ✓ | ✓ | ✓ | ✓ | ✓ | ✓ |
| Anxiety (severe) | ✓<br>(STAI-T) | ✓<br>(severe; ≤3y) | ✓<br>(severe psych.) | ✓<br>(strong fears/panic) | ✓ | ✓ |
| Severe mental illness (psychosis/bipolar etc.) | ✓<br>(any psych. disorder) | ✓<br>(psychosis; ≤3y) | ✓<br>(severe psych.) | ✓<br>(psychosis; FHx schizophrenia) | ✓ | ✓ |
| Pregnancy | ✓ | ✓ | — | ✓ | ✓ | ✓ |
| Unexplained loss of consciousness | ○ | ✓ | ○ | ○ | ✓ | ✓ |
| Neurological disease | ✓ | — | ✓ | ✓ | ○ | ✓ |
| Substance use | ✓<br>(≥14u/wk; <24h alcohol <1wk drugs) | — | ✓<br>(>3u <24h; any drugs <24h) | ✓<br>(psychotropics now/≤3mo) | ✓ | ✓<br>(<24h alcohol) |
| Psychiatric meds | ✓ | — | ✓ | ✓ | ✓ | ✓ |
| Sleep deprivation | — | — | ✓<br>poor sleep prev night | ✓<br>(acute/chronic) | ✓ | ✓ |
| Eye disease/uncorrected vision | ○<br>(uncorrected vision) | — | ✓<br>(eye disease) | — | ○ | ○ |
| Ear/vestibular (e.g., Ménière's) | — | — | ✓ | — | ○ | ○ |
| Cardiac issues or pacemaker | — | — | ✓ | — | ○ | ○ |
| Head trauma | ✓ | — | — | — | ✓ | ✓ |
| Age bounds | 21–60 | ≥18 | ≥18 | ≥18 | ≥18 | ≥18 |

All laboratories applied a minimum age criterion (18 or 21 years), reflecting higher PSE incidence in those aged 7–19 ^23^, as well as consent and safeguarding requirements for minors. Every protocol excluded a personal epilepsy history, particularly photosensitive subtypes, and most laboratories (A and B) also screened for first-degree family history. All laboratories assessed sensitivity to bright or flashing light (including migraine history or photophobia), and pregnancy was a common exclusion.

Despite convergence on core exclusions, laboratories differed across several domains:

Anxiety. All laboratories recognised potential discomfort in individuals with heightened anxiety, but screening approaches varied. Lab A used the State–Trait Anxiety Inventory, Trait version (STAI-T; ^48^) with an exclusion cut-off score of >50. Lab B excluded self-reported severe anxiety within the past 3 years, while Lab C used a broad "severe psychiatric condition" criterion. Lab D did not screen specifically for anxiety.

Psychiatric disorders. Laboratories A and C applied broad exclusions ("any psychiatric disorder"; "any severe psychiatric condition"). Lab D excluded psychosis or first-degree family history of schizophrenia, while Lab B excluded severe anxiety or psychosis within 3 years.

Alcohol and recreational drug use. Laboratories A, C, and D assessed recent use and instructed participants not to attend under the influence to reduce the risk of adverse reactions and preserve data quality. Laboratory B did not formally assess recent substance use.

Psychotropic medication use. Laboratories A, C, and D excluded current psychoactive medications (e.g., antipsychotics, anxiolytics), to avoid agents that may lower seizure threshold or alter SLS responsiveness. Laboratory B did not specify medication status.

Sleep deprivation. Laboratories C and D excluded recent sleep loss, whereas Labs A and B did not.

#### Current major medical issues

Laboratory A excluded active major medical conditions, including unstable hypertension, significant arrhythmias and multiple sclerosis. Photophobia has been reported in multiple sclerosis ^49^. This was a protocol-specific precaution; the present data do not establish these conditions as independent predictors of SLS-related events. Other laboratories did not apply this exclusion.

Traumatic brain injury (TBI). Laboratory A excluded any history of TBI, including concussion or post-concussive syndrome, given frequent post-TBI photophobia, headache, and sensory overload.

Unexplained loss of consciousness: Laboratory B explicitly screened for unexplained, repeated loss of consciousness; the other laboratories provided partial or qualified coverage of this history. Such episodes may indicate seizure, syncope or another condition requiring assessment.

Heart disease/pacemakers: Only Laboratory C explicitly excluded participants with heart disease or implanted cardiac pacemakers, including those without neuroimaging components, to maintain consistency in safety screening and minimise any potential cardiovascular risk.

Aphantasia: Laboratory C excluded aphantasia (inability to generate visual imagery voluntarily). This was a study-design choice to maintain comparable phenomenological reporting. Data on SLS-induced experiences in aphantasia remain limited ^50^.

### 3.4 Additional risk management strategies

Beyond screening, we note additional risk management strategies, identify common strategies across laboratories, and highlight areas of divergence.

First-aid training and on-site readiness. Most laboratories ensured that staff conducting studies had current first-aid training, including the recognition and management of seizures. In Laboratory C, not all staff were first-aid trained; however, a designated first-aid warden was always on call and immediately available during data collection.

#### Taster SLS exposure

All sites used graded exposure, including a brief taster session to confirm tolerability and identify potential adverse responses before the main experiment.

EEG pre-screening. In two studies, Laboratory D used a 10-minute EEG with intermittent photic stimulation for first-time SLS participants. Intermittent photic stimulation during EEG is an established clinical method for eliciting a PPR, but the incremental value of routine EEG screening in asymptomatic adults who have already undergone history-based screening is uncertain ^51^. A PPR may identify increased susceptibility, but a normal EEG does not establish that later exposure is risk-free and should not be treated as safety clearance. The present observational dataset did not compare screening strategies and cannot establish whether EEG screening, questionnaire screening or graded exposure is more effective.

### 3.5 Adverse responses to SLS

Across the laboratory reports, no seizures or syncopal episodes were reported. Contributors reported 20 non-serious events that interrupted or modified participation: 0 in Laboratory A, 7 in Laboratory B, 11 in Laboratory C and 2 in Laboratory D. Because participant deduplication, event definitions, monitoring procedures and follow-up differed across studies, we do not calculate a pooled prevalence or compare site-specific rates. Post hoc descriptive grouping identified anxiety or distress (n = 5), ocular discomfort or photophobia (n = 5), dizziness or vertigo (n = 3), headache or migraine (n = 3), and non-specific unpleasantness or early withdrawal (n = 4). No medical transfer was reported for the laboratory events. Event descriptions were supplied by contributing teams and were not independently verified against medical records; for online studies, follow-up was limited to information available to the original research teams.

## 4.0 Results: Commercial SLS Providers

### 4.1 Commercial providers screening procedures

Compared with laboratory protocols, commercial SLS screening procedures overlap on the core exclusions, mainly a history of epilepsy or PSE and age ≥18, with caution around migraine, photophobia and significant psychiatric histories (Table 1).

However, clear divergences from laboratory-based screening were observed. Provider A screened for migraine or photophobia, cardiac issues, sleep deprivation cut-offs, and neurodiversity-related sensory intolerance. Provider A also provided users with a detailed list of medications associated with increased seizure risk. In contrast, Provider B included core exclusions (e.g., epilepsy, pregnancy, recent alcohol/drugs, sleep deprivation, head injury), but did not formally assess photophobia or migraine, syncope history, cardiac or pacemaker status, or neurodiversity-related sensory needs.

### 4.2 Commercial providers adverse responses to SLS

Provider A. Across approximately 3.8 million provider-counted closed-eye sessions, nine events classified by the provider as requiring medical attention and as having probable or higher causal relation to SLS were reported through customer feedback and incident channels. The provider classified all nine as seizures. Non-serious reactions were not systematically recorded. Because reporting was passive, the completeness of case capture and follow-up is unknown; the count should not be interpreted as an incidence estimate.

Provider B. The provider reported more than 400,000 professional closed-eye sessions and five notable events through ad hoc feedback: one episode of transient memory loss after repeated use, one seizure at a public event, one prolonged trance-like unresponsive state, and two further seizures assessed by emergency services. The available records do not establish complete ascertainment, unique-user exposure, consistent causal assessment or systematic follow-up. We therefore report the five descriptions without estimating an event rate.

## 5.0 Discussion

Bringing these sources together provides a cross-setting account of reported adverse responses to SLS. Laboratory contributors reported 20 non-serious events and no seizures or syncope, whereas commercial providers separately reported nine and five medically important or otherwise notable events. Although these counts cannot establish incidence, absolute risk, causal attribution or screening effectiveness, they describe the events that were reported and identify what stronger evaluation requires: systematic surveillance, comparable exposure denominators and independent medical adjudication.

Building on this synthesis, we developed the 4SQ: a brief, plain-language, non-diagnostic checklist for structuring risk review and guiding proportionate precautions. It focuses on specific histories and sensitivities—including seizure history, previous visually provoked events, photophobia, migraine and sensory intolerance—rather than treating diagnostic labels as proxies for risk. Among the protocols and published guidance identified in this review, the 4SQ appears to be the most comprehensive publicly described SLS-specific pre-exposure checklist to date. What this establishes is breadth, not validity: it does not show that the 4SQ predicts adverse responses or reduces risk. These remain empirical questions. The next step is formal evaluation of its predictive performance, acceptability, and false-positive and false-negative rates.

### 5.1 Development of the Sussex Strobe Safety Screening Questionnaire (4SQ)

To translate the synthesis into practical questions, we considered each safety-relevant factor in relation to direct SLS evidence, clinical photosensitivity evidence and tolerability considerations. We then drafted plain-language questions and provisional follow-up actions. Where evidence was limited, recommendations were framed as precautionary rather than evidence of demonstrated risk reduction. The resulting 4SQ should be treated as a checklist for structured assessment, not as a validated diagnostic or predictive instrument.

#### 1) Age (minimum age & safeguarding)

##### Age and safeguarding

###### Rationale

Photosensitivity is most commonly identified in younger people, but it may persist or present after age 50 ^23,52^. Although the incidence and mortality of status epilepticus increase in later life ^53^, this epidemiological pattern does not establish increased photosensitivity and therefore does not justify an upper-age threshold for SLS. Age alone should not be used as a contraindication to SLS. **Screening:** Confirm that participants meet the study’s adult consent and safeguarding requirements. Assess neurological history, comorbidity, medication and capacity to communicate discomfort and stop exposure according to individual clinical context rather than chronological age.

###### Default decision

Retain age 18 as the lower boundary of this adult-use framework. Do not exclude or require clinical clearance solely on the basis of age; seek appropriate clinical input only when an individual’s history raises concern.

##### 2) Neurological history

##### PSE or seizure history (including first-degree family history)

###### Rationale

SLS can provoke epileptic seizures in susceptible individuals. Photosensitive seizures are heritable ^24^, and genetic susceptibility contributes broadly to epilepsy ^25^. A first-degree family history of photosensitive epilepsy should prompt more detailed assessment, although the available evidence does not quantify the incremental risk for an asymptomatic adult. Any personal history of unprovoked seizure indicates uncertainty about individual triggers. Given this diagnostic ambiguity, a conservative approach treats any prior seizure history as a contraindication.

###### Screening

Assess personal epilepsy or seizure history, first-degree family history and prior visually provoked events.

###### Default decision

Exclude participants with any personal seizure history. As a precaution, exclude or seek clinical input when a first-degree family history of photosensitive epilepsy is reported.

##### Migraine and photophobia

###### Rationale

Bright or flickering light is frequently reported as a migraine trigger, although some apparent triggering may reflect photophobia during the premonitory phase ^35,38^. Migraine, visual aura and photophobia are treated primarily as tolerability markers in this framework; the present data do not establish them as predictors of serious SLS events.

###### Screening

Assess migraine history, visual aura, photophobia and previous responses to bright or flashing light.

###### Default decision

Do not apply an automatic diagnosis-based exclusion. Defer or seek clinical advice where symptoms are active, severe or previously triggered by flashing light; otherwise consider lower intensity, shorter exposure and enhanced monitoring.

##### Traumatic brain injury (TBI), concussion, or head injury (past 12 months)

###### Rationale

Post-concussive symptoms can include photophobia, headache and dizziness ^35^. High-intensity SLS may therefore be less tolerable while such symptoms are active, but direct evidence is lacking.

###### Screening

Assess any head injury or TBI within the past 12 months and the presence of persistent post-concussive symptoms.

###### Default decision

Defer exposure while post-concussive symptoms are active or where the clinical history indicates unresolved seizure risk. Any fixed exclusion interval should be justified for the population and exposure context.

Unexplained loss of consciousness/episodes of altered awareness

###### Rationale

Unexplained episodes may indicate undiagnosed epilepsy (e.g., absence seizures) or other neurological instability. SLS could precipitate adverse events in such cases.

###### Screening

Assess any history of fainting, blackouts or brief unresponsiveness without a clear medical explanation.

###### Default decision

Exclude participants with unexplained loss of consciousness; defer participation pending medical evaluation.

1. 3) Neurodevelopmental conditions & sensory sensitivity

Autism spectrum disorder (ASD)

###### Rationale

Some autistic individuals experience marked sensory intolerance, although direct evidence on SLS-specific risk is lacking. The relevant screening target is the individual’s reported visual or sensory sensitivity, not the diagnosis itself ^40,41^.

###### Screening

Assess light sensitivity and sensory intolerance.

###### Default decision

Do not blanket-exclude. Defer exposure or use accommodations when marked photophobia or sensory intolerance is reported, or if the participant prefers not to proceed.

##### Attention-deficit/hyperactivity disorder (ADHD)

###### Rationale

An online survey found higher self-reported photophobia among adults with ADHD or ADHD symptoms than among respondents without ADHD ^42^. Sensory sensitivity, rather than the diagnostic label, is the relevant consideration.

###### Screening

Assess photophobia and sensory intolerance using brief, direct questions.

###### Default decision

Do not blanket-exclude. Defer exposure or use accommodations when marked photophobia or sensory intolerance is reported; otherwise, include.

#### 4) Psychiatric history

##### Anxiety disorders / high anxiety

###### Rationale

SLS can elevate arousal and alter perception, potentially precipitating panic or distress. However, evidence remains limited, warranting a conservative approach.

###### Screening

Use the STAI-T where feasible (suggested cut-off score > 50) or assess self-reported severe anxiety or panic history.

###### Default decision

Use elevated anxiety scores or severe symptoms to prompt contextual review rather than automatic exclusion.

##### Psychosis (current or recent)

###### Rationale

SLS-evoked visual phenomena may interact with positive psychotic symptoms, increasing distress or confusion. However, evidence is limited, justifying a conservative approach.

###### Screening

Assess current psychotic symptoms and any recent psychotic episodes. *Default decision:* Exclude those with current psychotic symptoms; consider inclusion only in stable remission with explicit clinical clearance.

#### 5) Other medical conditions

##### Pregnancy

###### Rationale

Direct evidence on SLS exposure during pregnancy is lacking. Physiological changes, altered drug metabolism, disrupted sleep, stress and fatigue may affect seizure control during pregnancy ^32^, supporting a precautionary approach.

###### Screening

Assess current pregnancy status.

###### Default decision

Exclude participants who are currently pregnant.

##### Heart disease / implanted cardiac devices / unstable medical conditions

###### Rationale

Direct evidence that cardiac disease or implanted devices increase SLS risk is lacking. Unstable cardiac or systemic illness may nevertheless warrant deferral because an acute event could complicate monitoring and response.

###### Screening

Assess history of arrhythmias, unstable angina, heart failure, implanted devices, uncontrolled hypertension, or active neurological or systemic disease.

###### Default decision

Exclude participants with unstable cardiac disease or uncontrolled serious medical conditions; seek appropriate clinical or device-specific advice where an implanted cardiac device is present.

#### 6) Substances & sleep

##### Alcohol or recreational drug use (recent)

###### Rationale

Recent intoxication might alter neural responsiveness (depending on substance), judgment, self-report, timely withdrawal, and tolerability.

###### Screening

Assess whether alcohol or recreational drugs were used recently and whether any current or residual effects are present (e.g., “high”, hangover, withdrawal, unusual fatigue, poor sleep).

###### Default decision

Exclude or reschedule participants reporting or exhibiting current or residual effects.

###### Operational note

Sites should pre-specify conservative wash-out periods by substance class (e.g., alcohol, cannabis, stimulants, MDMA, psychedelics).

##### Sleep deprivation (recent)

###### Rationale

Acute sleep loss can increase seizure susceptibility and may reduce a participant’s capacity to monitor and respond to distress ^26,27,30,31^.

###### Screening

Assess hours slept in the past 24 hours and recent acute restriction (e.g., <4 hours).

###### Default decision

Exclude or reschedule following acute sleep deprivation; proceed once sleep has normalised.

#### 7) Medications

##### Medication and withdrawal contexts

###### Rationale

Medication-associated seizure risk depends on the drug, dose and context and may be concentrated during toxicity, rapid dose change, withdrawal, interacting illness or combinations of pro-convulsant factors ^58,59^. The supplementary medication list is therefore provided as an awareness aid for researchers and organisers, not as prescribing guidance or a universal exclusion list.

###### Screening

Record current medication, recent initiation or dose change, withdrawal and any history of medication-associated seizures. Where necessary, eligibility decisions should be informed by a clinician or pharmacist. Participants should not discontinue or alter prescribed medication in order to take part.

###### Default decision

Defer participation or obtain appropriate clinical advice when there is a specific high-risk medication, toxicity, withdrawal, recent dose change or relevant seizure history. Stable medication use should be assessed in relation to the study population, protocol and clinical oversight rather than treated as an automatic exclusion.

We designed the Sussex Strobe Safety Screening Questionnaire (4SQ; Fig. 1) based on our review findings. The 4SQ operationalises these considerations into a consistent pre-exposure conversation covering neurological history, visually triggered symptoms, psychiatric stability, sensory tolerability, relevant medical conditions, substances, sleep and medication context. It has not yet been validated for sensitivity, specificity, predictive value or reduction of adverse events.

**Fig. 1.**
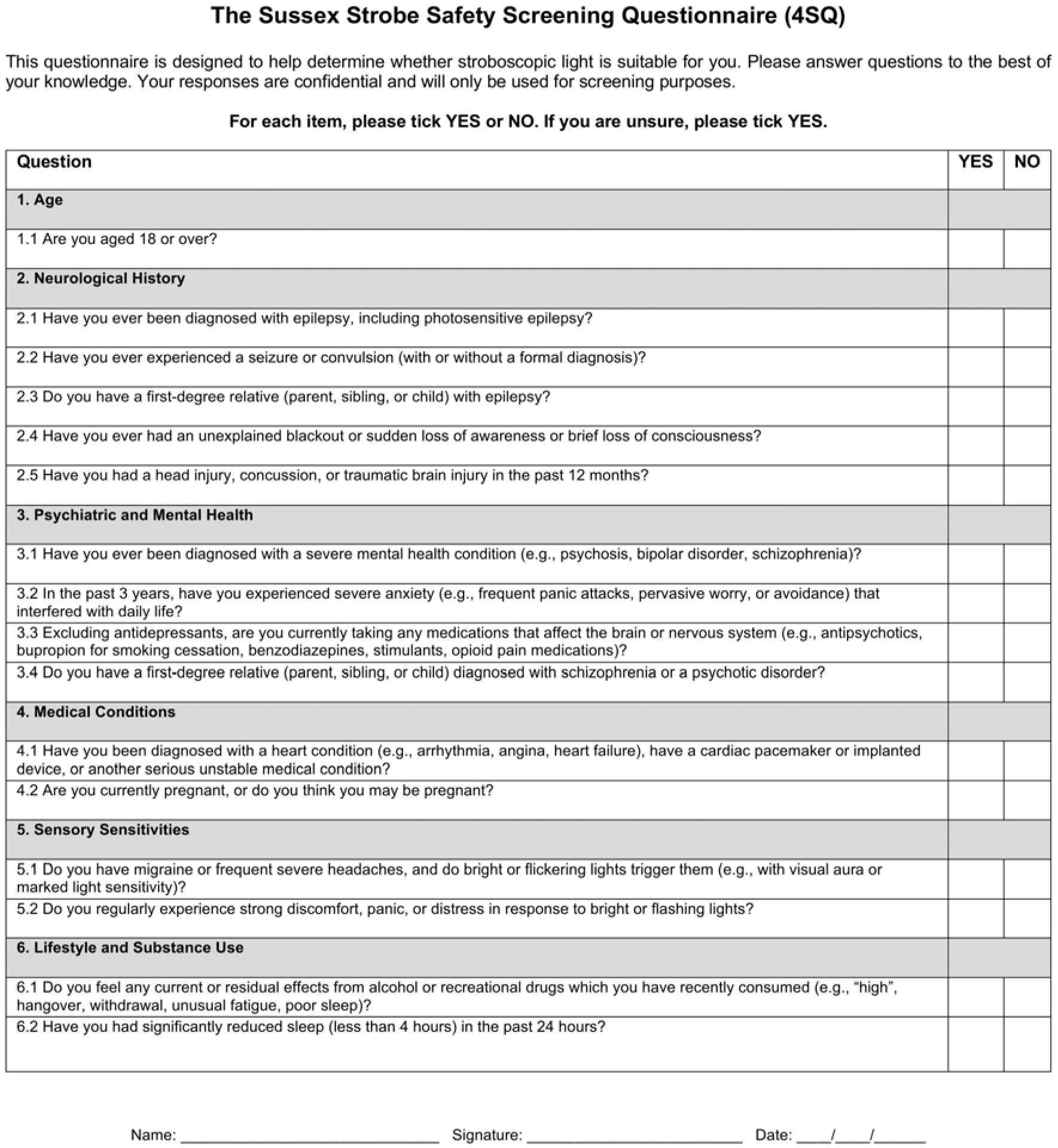
The Sussex Strobe Safety Screening Questionnaire (4SQ), a provisional, plain-language pre-exposure checklist for stroboscopic light stimulation (SLS). It covers neurological history, psychiatric and mental-health history, cardiovascular conditions, sensory sensitivities, and lifestyle or physiological factors. Responses are recorded as YES/NO. A YES response prompts further assessment and a context-specific decision, such as deferring exposure, seeking clinical advice, proceeding with caution or adding accommodations; it does not automatically establish ineligibility. The 4SQ is not a diagnostic instrument and has not yet undergone formal validation.

A ‘YES’ response to any item prompts further assessment and a predefined, context-specific eligibility decision (exclusion, inclusion with caution, or inclusion with accommodations), rather than automatic exclusion.

### 5.2 Practical risk mitigation for SLS

SLS risk management should combine appropriate participant information, context-specific screening, controlled exposure, a readily available stop procedure, trained staff and systematic event documentation. Common practices across the contributors included briefing, an explicit opt-out and a brief supervised familiarisation exposure. This familiarisation allows participants to experience the stimulus at conservative intensity, practise the stop procedure and identify immediate discomfort before the full session. It is retained as a tolerability measure intended to reduce avoidable discomfort and early distress. The present data did not compare this practice experimentally, and a tolerated familiarisation exposure does not constitute medical clearance or guarantee tolerance of longer, brighter or otherwise different stimulation.

In contexts where direct supervision is not possible, such as app-based or home-use SLS, additional safeguards are required. These include a simple and immediately accessible means to terminate stimulation (e.g., a clear “stop” control), automatic termination of SLS if sudden movement is detected where feasible, and user-adjustable brightness to enhance comfort and minimise minor side effects. Clear plain-language guidance should be provided on safe usage (e.g., avoiding use near water, in the bath, or at height). Robust incident-reporting mechanisms should be in place so that users can easily report adverse reactions. Following any report of a major incident, users should be guided towards appropriate medical attention, ideally under the oversight of medical professionals or designated safety officers.

Clinically, photosensitivity is assessed using intermittent photic stimulation during EEG. Provocation depends jointly on flash frequency, luminance and contrast, colour, stimulated visual-field area, viewing distance, and exposure duration. It also depends on whether the eyes are open or closed and on the act of eye closure itself. Current review evidence identifies risk across approximately 3–60 Hz, with greatest sensitivity often around 15–20 Hz, rather than establishing a single universally safe upper-frequency threshold ^24^. SLS protocols should document and control the full stimulus profile, minimise exposure to more provocative combinations, use gradual monitored exposure where appropriate, and avoid describing sub-15-Hz stimulation as intrinsically safe.

Photobiological safety cannot be inferred from luminous flux (lumens) alone because ocular hazard depends on spectral radiance or irradiance, source geometry, retinal image size and exposure duration. Closed-eye presentation should not be assumed to reduce seizure provocation: closing both eyes is not generally protective and can be more provocative in some photosensitive individuals ^24^. Neither blinking nor closed-eye presentation should be treated as a substitute for device-specific photobiological assessment. Protocols should measure relevant optical quantities at the participant position and document compliance with applicable guidance and standards, including the ICNIRP guidelines for incoherent visible and infrared radiation and IEC 62471 where relevant ^56,57^.

### 5.3 First-episode risk without screening: benchmarks and context

A residual possibility of a first visually triggered seizure remains because some susceptible individuals may have no previous diagnosis or recognised visually triggered event. However, we cannot quantify that probability or the proportion removed by screening. Evidence from occupational EEG testing and public events is useful as context only because outcomes, populations and exposures differ substantially from closed-eye SLS.

In UK aircrew candidates, 44 of 13,658 applicants showed epileptiform activity only during photic stimulation, while one photically induced convulsion was reported ^54^. In 5,893 Danish pilot applicants, 128 intermittent-photic-stimulation EEG abnormalities and four IPS-related seizures were reported ^55^. At electronic dance-music festivals, 39 seizure episodes were recorded across 400,343 cumulative attendances, with more episodes at night than during daylight events ^46^. These observations mix EEG responses with clinical seizures, use attendance rather than unique-person denominators, and differ in stimulus and co-exposures.

We do not pool these studies or interpret them as estimates of first-episode SLS risk. A PPR is not equivalent to a seizure, cumulative attendance is not a unique-person denominator, and the festival comparison is vulnerable to differences in sleep, substances, temperature, crowding and event duration. The studies instead demonstrate why further SLS research should distinguish EEG photosensitivity, visually triggered seizures and non-epileptic adverse responses.

Prior symptom-free exposure to flashing light may contribute useful history, but it does not establish safety under a different frequency, brightness, field size, colour, duration or eye-state condition. It should prompt contextual follow-up rather than operate as a clearance test.

Clear participant information, an explicit opt-out, readily available stopping procedures, monitored initial exposure and trained staff are proportionate precautions. Their effectiveness, and the incremental value of each screening question, require formal evaluation using predefined exposure denominators and systematic follow-up.

### 5.4 Limitations

This synthesis has several limitations. The contributors were selected as they had substantial experience delivering SLS; they were not sampled systematically, and reports were retrospective. Most laboratory studies involved young adult, predominantly university or student samples, apart from the broader depression studies; commercial user characteristics were less completely described. Participants were not counted more than once within an individual study, but some may have participated in more than one study because cross-study deduplication was unavailable. Study populations, devices, frequencies, brightness metrics, exposure durations, screening criteria, event definitions and follow-up procedures varied. This diversity helped identify safety considerations that recurred across delivery contexts, supporting guidance that is not tied to any single device or protocol. However, protocol heterogeneity, passive event reporting, non-comparable exposure denominators and the absence of verified links between reported events and individual users or sessions prevent pooled incidence estimation and comparisons of the relative safety of individual protocols. The provider-specific ratios should therefore be interpreted as descriptive reporting frequencies, not as incidence estimates or formal lower bounds. The absence of a reported event is not evidence that no event occurred. These limitations preclude conclusions about absolute risk, causal comparison of protocols or the effectiveness of screening. The 4SQ is a provisional checklist whose predictive performance requires formal evaluation.

## 6.0 Conclusion

This synthesis provides a cross-setting account of current SLS screening practice and reported adverse responses across heterogeneous laboratory and commercial settings. Laboratory contributors reported no seizures or syncope, whereas commercial providers separately reported nine and five medically important or otherwise notable events through passive channels. These observations describe reported events but cannot establish incidence, absolute risk or screening effectiveness. The 4SQ consolidates the recurrent safety considerations identified across these settings into a single, non-diagnostic framework for structured pre-exposure review, although its predictive performance remains to be established. Together, the synthesis and 4SQ provide a practical foundation for more consistent risk management and for formal evaluation of screening performance and adverse-event burden using harmonised definitions, unique-person and session denominators, and systematic follow-up.

## Data Availability

All data produced in the present study are available upon reasonable request to the authors

## Acknowledgements

The authors thank Collective Act Ltd and its Director, Jennifer Crook, for delivering the Dreamachine programme and collecting source data that contributed to this synthesis. The authors also thank Devraj Joshi (Whatever Together) and Cecelia Schwartzman for helpful discussions.

## Data Availability

The aggregate reported-event data, survey instrument, 4SQ and accompanying coding documentation are available at https://osf.io/nwkev/overview?view_only=c9979a478c734e7a9e46e50b5bed5c3a. The authors received no identifiable participant-level data. Source records supplied by laboratories and commercial providers are not publicly shared where institutional, privacy or commercial restrictions apply. The repository documents the provenance and limitations of the aggregate records used in the analysis.

---

**S1 Table.** Available study and portfolio-level characteristics of contributing laboratory datasets and adverse-event ascertainment.

**S1 Text.** Medications reported to lower seizure threshold: awareness aid and interpretive limitations.

