## Supplemental Table S1 for "Stroboscopic Light Stimulation Safety Within and Beyond Laboratory Settings: Observational Evidence and Practical Guidance"

**S1 Table. Available study and portfolio-level characteristics of the contributing laboratory datasets, together with information on adverse-event ascertainment.**

*The table reflects the information available to the authors. Some contributor records group multiple related studies or provide only portfolio-level totals; these are identified as such rather than disaggregated without confirmation from the source. NR indicates information not reported in the available source records. All values are descriptive, with unresolved discrepancies stated explicitly. Event counts refer to reported events; they are not incidence estimates.*

| **Study** | **Purpose / population / setting** | **Device / eye state** | **Frequency / sequence** | **Exposure / sessions** | **Exposed N** | **Screening / AE ascertainment** | **Reported events / follow-up / source note** |
| --- | --- | --- | --- | --- | --- | --- | --- |
| Laboratory A; A-01 | Laboratory study; mostly university students; mean age 23.94 years; laboratory | Custom Lumenate research lamp; 12 white LEDs; eyes closed | 10 Hz or constant light | NR; sessions/person: NR | 20 (reported) | Extensive neurological, psychiatric, migraine and medication exclusions; familiarisation: NR; ascertainment: Direct contributor survey; calamity protocol | Non-serious: 0; serious: 0; follow-up: next-day follow-up specified if an event occurred. The contributor report listed N = 20; a separate exposed-participant count was not available. |
| Laboratory B; B-WP1 | Parameter-optimisation study; adults reporting depressive symptoms; laboratory | Roxiva; eyes closed | Wide parameter range plus dynamic sequence | Eleven sessions with 30-second parameter sections; sessions/person: 11 | 31 | 4SQ-style screening; 14/53 screened out; familiarisation: Yes; ascertainment: Tolerability scale and researcher observation | Non-serious: 1; serious: 0; follow-up: NR. One participant stopped after tolerability score 8. |
| Laboratory B; B-WP2 | Randomised repeated-session study; adults reporting depressive symptoms; laboratory | Roxiva; eyes closed | Dynamic intervention and lower-phenomenology control | NR; sessions/person: 4 | NR | Safety screening before enrolment; familiarisation: NR; ascertainment: session side-effect measures and researcher observation | Non-serious: NR; serious: 0; follow-up: NR. The contributor workbook reported a combined N = 105 across the two depression studies; a study-specific exposed N and any overlap were not reported. |
| Laboratory B; B-INT | Repeated-session study; neurotypical adults; laboratory | Roxiva; eyes closed | Dynamic sequence | NR; sessions/person: NR | 33 | 24/98 failed safety screening; additional exclusions; familiarisation: NR; ascertainment: NR | Non-serious: 0; serious: 0; follow-up: NR. Possible participation in other Laboratory B studies could not be assessed from the available records. |
| Laboratory B; B-EEG | EEG strobe study; Healthy adults; Laboratory | Lucia; eyes closed | 3, 8 and 15 Hz plus 8 Hz aperiodic | NR; sessions/person: NR | 36 | Epilepsy, migraine, light sensitivity and STAI >50 exclusions; familiarisation: Yes; ascertainment: Researcher observation and withdrawal record | Non-serious: 1; serious: 0; follow-up: NR. One participant withdrew after taster because they did not enjoy it. |
| Laboratory B; B-MSC | Laboratory study; adults aged 18–35; laboratory | Research strobe; eyes closed | NR | NR; sessions/person: NR | 34 | Screening: NR; familiarisation: NR; ascertainment: researcher observation | Non-serious: 1; serious: 0; follow-up: NR. One participant stopped after the second trial. |
| Laboratory B; B-ALPHA | Alpha-suppression study; Healthy adults; Laboratory | Lucia; eyes closed | NR | NR; sessions/person: NR | 29 | No safety or STAI exclusions reported; familiarisation: NR; ascertainment: NR | Non-serious: 0; serious: 0; follow-up: NR. |
| Laboratory B; B-IMAGERY | Imagery study (main experiment); healthy adults; laboratory | Lucia; eyes closed | Dynamic sequence | NR; sessions/person: NR | 26 | Screening: NR; familiarisation: NR; ascertainment: NR | Non-serious: 0; serious: 0; follow-up: NR. The contributor workbook referred to eight related studies, but detailed records were available only for this main experiment. |
| Laboratory B; B-LEM | Two-experiment perceptual-task study; healthy adults; laboratory | Bespoke stroboscope; eyes closed | NR | NR; sessions/person: NR | 33/39* | No exclusions reported; familiarisation: NR; ascertainment: Researcher withdrawal records | Non-serious: 2 as coded in the contributor workbook; serious: 0; follow-up: NR. The sample-size discrepancy was unresolved. Additional non-completions were described but could not be classified against the survey event definition from the available records. |
| Laboratory B; B-TR1 | Phenomenology study 1; adults aged 18–35; laboratory | Lucia No. 3; eyes closed | Includes 3 Hz | NR; sessions/person: NR | NR | Earlier site screening form; familiarisation: NR; ascertainment: researcher observation | Non-serious: 1; serious: 0; follow-up: NR. One of four studies with a combined reported N = 159; study-specific N was not reported. |
| Laboratory B; B-TR2 | Phenomenology study 2; adults aged 18–35; laboratory | Roxiva; eyes closed | NR | NR; sessions/person: NR | NR | Current site screening form; familiarisation: NR; ascertainment: researcher observation | Non-serious: 0; serious: 0; follow-up: NR. One of four studies with a combined reported N = 159; study-specific N was not reported. |
| Laboratory B; B-TR3 | Phenomenology study 3; adults aged 18–35; laboratory | Roxiva; eyes closed | NR | NR; sessions/person: NR | NR | Current site screening form; familiarisation: NR; ascertainment: researcher observation | Non-serious: 0; serious: 0; follow-up: NR. One of four studies with a combined reported N = 159; study-specific N was not reported. |
| Laboratory B; B-TR4 | Phenomenology study 4; adults aged 18–35; laboratory | Roxiva; eyes closed | Includes 11 Hz | NR; sessions/person: NR | NR | Current site screening form; familiarisation: yes; ascertainment: researcher observation | Non-serious: 1; serious: 0; follow-up: NR. One of four studies with a combined reported N = 159; study-specific N was not reported. |
| Laboratory C; C-01 | MEG study; Healthy adults; Laboratory/MEG | Epson EB-G7400U projector; eyes closed | 15 Hz and constant light | NR; sessions/person: NR | 30 | Site safety protocol; familiarisation: NR; ascertainment: Researcher observation | Non-serious: 2; serious: 0; follow-up: Short-term symptoms recorded. NR |
| Laboratory C; C-02 | Laboratory/online screen flicker; Healthy adults; Mixed lab/online | Participant or laboratory PC; eyes closed | 15 Hz and 0.1 Hz | NR; sessions/person: NR | 140 | Site safety protocol; familiarisation: Yes; ascertainment: Lab observation or online discontinuation report | Non-serious: 2; serious: 0; follow-up: Unknown. NR |
| Laboratory C; C-03 | Online screen-flicker study; online participants; online | Participant PC/laptop; eyes closed | 15 Hz and 0.1 Hz | NR; sessions/person: NR | 90 | Site safety protocol; familiarisation: NR; ascertainment: Online non-completion report | Non-serious: 2; serious: 0; follow-up: Unknown. NR |
| Laboratory C; C-04 | Online guided-meditation study; Online participants; Online | Participant PC/laptop; eyes closed | 10 Hz and 0.1 Hz | NR; sessions/person: NR | NR | Site safety protocol; familiarisation: NR; ascertainment: Online non-completion report | Non-serious: 5; serious: 0; follow-up: unknown. The event narrative reported N = 168 and the demographic description N = 164. |
| Laboratory C; C-05 | Lab screen flicker; NR; Laboratory | Lab PC; eyes open or closed | 15 Hz | NR; sessions/person: NR | NR | Site safety protocol; familiarisation: NR; ascertainment: NR | Non-serious: 0; serious: 0; follow-up: NR. Participant N and ascertainment details were not reported. |
| Laboratory C; C-06 | Colour screen flicker; NR; Laboratory | Lab PC; eyes closed | 15 Hz | NR; sessions/person: NR | NR | Site safety protocol; familiarisation: NR; ascertainment: NR | Non-serious: 0; serious: 0; follow-up: NR. Participant N and ascertainment details were not reported. |
| Laboratory D; D-01 | Laboratory study 1; healthy adults; laboratory | Lucia No. 3 / research lamp; eyes closed | 1-40 Hz | NR; sessions/person: NR | 24 | Site screening plus prior tolerance/EEG option; familiarisation: NR; ascertainment: Researcher observation | Non-serious: NR; serious: 0; follow-up: NR. Laboratory D reported two non-serious events across five studies; study-level allocation was not reported. |
| Laboratory D; D-02 | Laboratory study 2; healthy adults; laboratory/fMRI | Lucia No. 3 / research lamp; eyes closed | 1-40 Hz | NR; sessions/person: NR | 24 | Site screening plus prior tolerance/EEG option; familiarisation: NR; ascertainment: Researcher observation | Non-serious: NR; serious: 0; follow-up: NR. Study-level event allocation was not reported. |
| Laboratory D; D-03 | Laboratory study 3; healthy adults; laboratory | Lucia No. 3 / research lamp; eyes closed | 1-40 Hz | NR; sessions/person: NR | 20/21* | Site screening plus prior tolerance/EEG option; familiarisation: NR; ascertainment: Researcher observation | Non-serious: NR; serious: 0; follow-up: NR. Study-level event allocation was not reported. |
| Laboratory D; D-04 | Laboratory study 4; healthy adults; laboratory | Lucia No. 3 / research lamp; eyes closed | 1-40 Hz | NR; sessions/person: NR | 20 | Site screening plus prior tolerance/EEG option; familiarisation: NR; ascertainment: Researcher observation | Non-serious: NR; serious: 0; follow-up: NR. Study-level event allocation was not reported. |
| Laboratory D; D-05 | Laboratory study 5; healthy adults; laboratory | Lucia No. 3 / research lamp; eyes closed | 1-40 Hz | NR; sessions/person: NR | 20 | Site screening plus prior tolerance/EEG option; familiarisation: NR; ascertainment: Researcher observation | Non-serious: NR; serious: 0; follow-up: NR. Study-level event allocation was not reported. |
