## Supplemental Text S1 for "Stroboscopic Light Stimulation Safety Within and Beyond Laboratory Settings: Observational Evidence and Practical Guidance"

**S1 Text. Medications reported to lower seizure threshold: awareness aid and interpretive limitations**

***Interpretive note.*** This list is an awareness aid for researchers and organisers, not prescribing guidance or a universal exclusion list. Medication-associated seizure risk depends on the drug, dose and context. Eligibility decisions should therefore be specific to the protocol and, where necessary, informed by a clinician or pharmacist. Participants should not discontinue or alter prescribed medication in order to take part. See supplementary references 1,2.

The following list summarises medications and medication-related contexts reported to lower seizure threshold:

**Antimicrobials**

- β-lactams: penicillins, cephalosporins, carbapenems (dose-related risk; carbapenems higher relative risk; also rapid, marked reduction of valproate levels with carbapenems).
- Isoniazid (mechanism via pyridoxal-5-phosphate/GABA; pyridoxine reverses toxicity).
- Fluoroquinolones (e.g., ciprofloxacin, ofloxacin, levofloxacin; rare but reported; GABAA antagonism).

**Antimalarials**

- Mefloquine, chloroquine (seizures reported in people with and without epilepsy).

**Analgesics**

- Opioids (class effect is context- and dose-dependent; tramadol, buprenorphine and pethidine/meperidine most consistently implicated; others possible under certain conditions).
- NSAIDs: aspirin, diclofenac, indometacin (dose-dependent; uncommon clinically). Mefenamic acid can be pro-convulsant at toxic doses. Ibuprofen and paracetamol are not implicated.

**Methylxanthines (respiratory)**

- Theophylline, aminophylline (adenosine A1 antagonism; can cause difficult-to-treat seizures in toxicity).

**Antipsychotics**

- Clozapine (strongest signal; clinical seizures ~3–6%).
- Chlorpromazine and other phenothiazines (other antipsychotics have lower but present risk).
- Haloperidol, benperidol, droperidol, melperone, azaperone (butyrophenones)

**Antidepressants / other monoaminergic agents**

- Venlafaxine and mirtazapine (rare, usually dose-related pro-convulsant effects, most often reported in overdose or at higher therapeutic doses).

**Antiepileptics (paradoxically)**

- Benzodiazepine withdrawal (withdrawal lowers threshold).
- Abrupt withdrawal of topiramate, lamotrigine, gabapentin or pregabalin can precipitate seizures.
- Carbamazepine (can exacerbate some generalised/myoclonic epilepsies).

**Immunosuppressant / Antineoplastic**

- Cyclosporine (high epileptogenic potential in review).
- Chlorambucil (intermediate potential).

**Radiology contrast**

- Iodinated contrast media (low epileptogenic potential; rare events; risk affected by CNS comorbidity).

**Smoking cessation**

- Bupropion (clear, dose-dependent risk at therapeutic use for smoking cessation).

**Other**

- Interferon-α (minimal/inconclusive pro-convulsant potential).
